# Blockade of Interleukin Seventeen (IL-17A) with Secukinumab in Hospitalized COVID-19 patients – the BISHOP study

**DOI:** 10.1101/2021.07.21.21260963

**Authors:** Gustavo Gomes Resende, Ricardo da Cruz Lage, Samara Quadros Lobê, Amanda Fonseca Medeiros, Alessandra Dias Costa e Silva, Antônio Tolentino Nogueira Sá, Argenil José de Assis Oliveira, Denise Sousa, Henrique Cerqueira Guimarães, Isabella Coelho Gomes, Renan Pedra Souza, Renato Santana Aguiar, Roberto Tunala, Francisco Forestiero, Julio Silvio Souza Bueno Filho, Mauro Martins Teixeira

## Abstract

**Background:** Patients with severe COVID-19 seem to have a compromised antiviral response and hyperinflammation. Neutrophils are critical players in COVID-19 pathogenesis. IL-17A plays a major role in protection against extracellular pathogens and neutrophil attraction and activation. We hypothesized that secukinumab, an anti-IL17A monoclonal antibody, could mitigate the deleterious hyperinflammation in COVID-19.

**Methods:** BISHOP was an open-label, single-center, phase-II controlled trial. Fifty adults hospitalized Covid-19 patients, confirmed by a positive SARS-CoV-2 RT-PCR, were randomized 1:1 to receive 300mg of secukinumab subcutaneously at day-0 (group A) plus standard of care (SoC: antiviral drugs, antimicrobials, corticosteroids, and/or anticoagulants) or SoC alone (group B). A second dose of 300mg of secukinumab could be administered on day-7, according to staff judgment. The primary endpoint was ventilator-free days at day-28 (VFD-28). Secondary efficacy and safety outcomes were also explored.

**Findings:** An intention-to-treat analysis showed no difference in VFD-28: 23.7 (95%CI 19.6-27.8) in group A vs. 23.8 (19.9-27.6) in group B, p=0.62; There was also no difference in hospitalization time, intensive care unit demand, the incidence of circulatory shock, acute kidney injury, fungal or bacterial co-infections, and severe adverse events. Pulmonary thromboembolism was less frequent in group A (4.2% vs. 26.2% p=0.04). There was one death in each group. Viral clearance, defined by the viral load fold change (2-ΔΔCT) in upper airways, between day-0 and day-7, was also similar: 0.17 (0.05-0.56) in group A vs. 0.24 (0.10-0.57) in group B.

**Interpretation:** The efficacy of secukinumab in the treatment of Covid19 was not demonstrated. No difference between groups in adverse events and no unexpected events were observed.

**Funding:** Novartis Brazil supported this research providing expert input in the development of the project, drug supply, data management, and monitoring.

## 1. INTRODUCTION

The new coronavirus (SARS-CoV-2) pandemic, which started in late 2019, has spread with impressive speed across the planet and has threatened all health systems due to the considerable proportion of patients requiring inpatient treatment, including intensive care and mechanical ventilation ^(1)^. The most severe patients, who progress to acute respiratory distress syndrome (ARDS), seem to combine a poor antiviral response and systemic hyperinflammation ^(2-5)^. Their laboratory findings (liver dysfunction, hyperferritinemia, disseminated intravascular coagulation, high levels of D-dimer, and C-reactive protein, along with cytopenias) and cytokine responses resemble those of other Hyperferritinemic Syndromes ^(6)^ such as the macrophage activation syndrome (MAS), adult-onset Still’s disease (AOSD), septic shock and catastrophic antiphospholipid syndrome (CAPS) ^(7, 8)^.

It has been suggested that neutrophils are critical players in COVID-19 pathogenesis. Indeed, COVID-19 severity has been associated with neutrophilia, increased neutrophil-to-lymphocyte ratio (NLR), and aberrant neutrophil activation ^(9)^. Moreover, neutrophils infiltrate the lungs where diffuse alveolar damage and microvascular thrombosis have been observed ^(10)^. IL-17 is a cytokine contributing to the pathogenesis of several immune-mediated diseases, including spondyloarthritis and psoriasis ^(11, 12)^. IL-17 drives neutrophil accumulation in these diseases and can up-regulate several cytokines, including CXCL8, TGF-β, IL-1β, TNFα, and IL-6 ^(13-15)^. In the context of COVID-19, some studies suggest that IL-17 may contribute to the pathogenesis of severe COVID-19. It was already showed that Th17 count and IL-17 levels were increased in critically ill COVID-19 patients compared to healthy donors. In addition, the ratio of Th17/Treg cells, RORγt/FoxP3, and IL-17/IL-10 were considerably enhanced in patients who evolved to death compared to recovered cases ^(16)^. Higher concentrations of IL-17A in blood have been associated with a more severe clinical course in patients with COVID-19. An expansion of tissue-resident memory-like TH17 cells (TRM17 cells) has been observed in the lungs even after viral clearance ^(17)^. Furthermore, treatment with an IL-17RA antibody decreased inflammation in the lungs of mice infected with SARS-CoV-2 pseudovirus expressing ORF8 virus protein^(18)^. Although these findings, a direct demonstration of the relevance of IL-17 in the pathogenesis of COVID-19 is lacking.

Secukinumab is a human IgG1κ monoclonal antibody that binds to and neutralizes IL-17A. It is currently approved in many countries to treat psoriasis, ankylosing spondylitis, non-radiographic axial spondyloarthritis, and psoriatic arthritis. We hypothesized that secukinumab could be used in patients with respiratory failure due to COVID-19 and report here the results of a phase 2 open-label randomized controlled trial to evaluate the effects of secukinumab in hospitalized adult COVID-19 patients.

## 2. METHODS

### Study design and participants

BISHOP (**<u>B</u>**lockade of **I**nterleukin **<u>S</u>**eventeen in **<u>H</u>**ospitalized c**<u>O</u>**vid-19 **<u>P</u>**atients) is an investigator-initiated, open-label, single-center, phase-II controlled trial. The study was performed in Hospital Risoleta Tolentino Neves (HRTN), referral center for treatment of COVID-19 in northern region of Belo Horizonte city in Brazil. Adult patients (age ≥ 18 years) admitted to the hospital were eligible for the study if they had SARS-CoV-2 infection confirmed by RT-PCR of nasopharyngeal swab and severe acute respiratory syndrome (SARS), according to the Brazilian Ministry of Health criteria (dyspnea / respiratory discomfort OR persistent chest pressure OR oxigen saturation less than 95% in room air OR cyanosis of lips or face) ^(19)^. Patients were included from September 1^st^ to December 29^th^, 2020.

The exclusion criteria were functional classes III and IV of congestive heart failure (CHF) or chronic obstructive pulmonary disease (COPD), stages 4 and 5 of chronic kidney disease (GFR <30 mL/min), diabetic ketoacidosis, known history of HIV/AIDS infection or chronic or acute HBV or HCV infection, unrestrained bacterial or fungal co-infections (defined by hemodynamic instability, prior 48h of antimicrobial covering, or at the discretion of the study’s medical coordinator), active tuberculosis, history of malignancy in the last year, current use (or in the previous 15 days) of other immunosuppressants, baseline neutrophils count ≤1000/mm3, pregnancy or breastfeeding.

### Randomization and procedures

Enrolled patients were sequentially subjected to block randomization to receive 300 mg of secukinumab subcutaneously at day-0 plus standard of care (SoC), hereinafter called group A, or SoC alone (group B) in a 1:1 ratio. A second dose of 300 mg of secukinumab could be administered at day 7, according to the judgment of the attending medical team and if theall enrollment criteria still remained met. Of note, secukinumab dose regimen in COVID-19 patients has not been assessed previously. The standard of care in Hospital Risoleta Tolentino Neves, available to all patients, regardless of their allocation, during all the time of study execution, included antimicrobials (azithromycin and a beta-lactam, at the time of admission and until bacterial infection of the lungs could be excluded), corticosteroids (dexamethasone, 6mg daily, during ten days), and prophylactic anticoagulants (enoxaparin 40mg q12hr, except if thromboembolism was diagnosed, in which case full anticoagulation was performed). Other immunobiological agents, such as anti-IL-6R / anti-IL1β / anti-TNF-α, was not allowed. Patients, after hospital discharge, were monitored by phone calls and messages until day 28.

### Outcomes

The primary endpoint was ventilator-free days at day 28 (VFD-28). VFD-28 is defined as (28 - x), where x is the number of days spent receiving mechanical ventilation. This outcome focuses on the duration of need for mechanical ventilation while simultaneously taking mortality into consideration because it assigns a score of zero if the patient dies before 28 days ^(20)^. Secondary efficacy and safety outcomes also included (all assessed within the time frame of 28 days): mortality rate; length of hospitalization; need and length of critical care in the ICU; World Health Organization (WHO) Ordinal Scale for Clinical Improvement; serious adverse events (any undesirable event associated at least with prolongation of hospitalization), secondary bacterial and fungal infections (culture-confirmed); and the incidence of acute renal failure (KDIGO III), circulatory shock or need for vasopressors, pulmonary thromboembolism (PTE) and reactions at the injection site. The effects of secukinumab on viral clearance were explored by longitudinal sampling and RT-PCR for SARS-CoV-2 in the upper airway swab. Data were collected using the Research Electronic Data Capture (REDCap) database (https://www.project-redcap.org).

### Statistical analysis

An overall type I error (2-sided) probability of 5% was used. At the time of the study design, there was a scarcity of published data on studies evaluating the effects of anti-inflammatory therapies in patients with COVID-19. A review and meta-analysis on the use of corticosteroids in ARDS found a mean difference in VFD-28 of 5.8 days (SD 7.8), favoring its use compared to placebo ^(21)^. The mean length of mechanical ventilation in critically ill patients with COVID-19 reported was 9.8 days (SD 4.4), and the mean calculated VFD-28 was 9.4 (SD 6.7) ^(22)^. The minimum sample size to achieve 80% power to detect an increase of 5.5 days in the VFD-28 was 25 patients per group.

Fisher’s exact test was used to evaluate frequencies among the two groups in the assessment of categorical data. For continuous numerical variables, the Mann-Whitney test was used to test differences in population distributions. A per-protocol (PP) analysis was used for all outcomes. Additionally, an intention-to-treat (ITT) analysis with covariable correction imputation method for missing data was done only for the primary endpoint.

The study was approved by the National Research Ethics Commission – CONEP (approval number 4.100.115, on June 20th, 2020) and was registered at the Brazilian Registry of Clinical Trials – ReBEC (RBR-5vpyh4, on June 6^th^, 2020). The informed consent form (ICF) was applied to all patients or their legal guardians for patients unable due to their medical condition.

## 3. RESULTS

Fifty patients were enrolled. The two randomized groups were homogenous in parameters of age, sex, and comorbidities (Table 1). Even though there were more systemic arterial hypertension, obesity/overweight, and cardiopathy in group A (treated patients) and more male patients in group B (control patients), none of these differences reached statistical significance. The mean (SD) age at study entry was 55 (11) years-old (YO) in group A and 52 (14) YO in group B. The mean (SD) time since onset of symptoms to randomization was 9.9 (2.7) days in group A and 9.5 (3.0) in group B.

**Table 1.** Baseline characteristics of patients randomized to secukinumab + standard of care (group A) or stand of care alone (group B).

| Group | A (SEC + SoC)<br>n=25 | B (SoC alone)<br>n=25 | p value |
| --- | --- | --- | --- |
| **Demographic characteristics** |  |  |  |
| Sex (male), n (%) | 10 (40) | 16 (64) | 0.16 |
| Age, median (IQR) | 54 (45-65) | 54 (40-63) | 0.37 |
| **Comorbidities, n (%)** |  |  |  |
| Systemic arterial hypertension | 13 (52) | 11 (44) | 0.78 |
| Diabetes mellitus | 8 (32) | 9 (36) | 1.00 |
| Obesity and overweight | 15 (60) | 9 (36) | 0.16 |
| Cardiopathy | 3 (12) | 1 (4) | 0.61 |
| Pneumopathy | 2 (8) | 3 (12) | 1.00 |
| N° of comorbidities/patient | 1.6 | 1.2 | 0.75 |
| **Clinical and laboratory measures, median (IQR)** |  |  |  |
| Time from symptoms onset to randomization (days) | 10 (8-12) | 9 (8-11) | 0.62 |
| Mean arterial pressure (mmHg) | 97 (93-100) | 94 (90-102) | 0.18 |
| Hemoglobin (g/dL) | 12.7 (12.1-14.3) | 13.4 (12.5-13.9) | 0.22 |
| White blood cells (10<sup>9</sup>/L) | 8.5 (7.6-9.8) | 8.9 (7.3-11.6) | 0.27 |
| Neutrophils (10<sup>9</sup>/L) | 6.8 (5.9-8.3) | 7.8 (5.8-9.3) | 0.34 |
| Lymphocytes (10<sup>9</sup>/L) | 1.3 (0.7-1.9) | 1.2 (0.9-1.5) | 0.68 |
| Neutrophil/lymphocyte ratio | 5.2 (3.6-10.5) | 6.5 (4.5-9.1) | 0.44 |
| Platelets (10<sup>9</sup>/L) | 251 (195-297) | 220 (171-301) | 0.32 |
| Creatinine (mg/dL) | 0.70 (0.70-1.0) | 0.75 (0.70-0.90) | 0.76 |
| Lactate dehydrogenase (U/L) | 331 (296-451) | 392 (324-499) | 0.15 |
| Ferritin (ng/mL) | 659 (344-1296) | 846 (428-1421) | 0.54 |
| C-reactive protein (mg/L) | 43 (24-77) | 63 (44-123) | 0.11 |
| Fibrinogen (mg/dL) | 461 (436-559) | 476 (442-527) | 0.57 |
| Troponin (ng/mL) | 3.5 (1.8-8.9) | 5.1 (3.3-14) | 0.22 |
| D-dimer (mcg/mL) | 0.6 (0.3-1.3) | 0.9 (0.4-2.3) | 0.21 |
| PaO2/FiO2 ratio | 297 (216-384) | 258 (174-339) | 0.07 |
| SOFA score | 2.0 (1.0-3.0) | 3.0 (1.0-3.0) | 0.10 |
| WHO scale | 4 (4-4) | 4 (4-4) | 0.48 |

IQR, interquartile range; PaO2/FiO2 ratio, ratio of arterial oxygen partial pressure (in mmHg) to fractional inspired oxygen; SOFA score, Sequential Organ Failure Assessment Score for sepsis; WHO scale, World Health Organization Ordinal Scale for Clinical Improvement.

During the trial, two patients were excluded, both from group B. One of them withdrew consent, alleging intolerable discomfort with sequential blood and nasopharyngeal swab collection. A second one was excluded due to a protocol deviation (only after enrolment, the patient was found to be receiving post-transplant immunosuppression).

Ten patients from group A were still hospitalized at day 7, and a second dose of 300 mg secukinumab was administered to seven of them. The remaining three patients did not receive the second dose, because two of them no more fulffilled SARS criteria and the and the third one was under septic shock.

The primary endpoint was defined as the number of ventilator-free days at day 28 (VFD-28). As seen in Table 2, there was no difference in VFD-28 between groups (23.7 days in group A and 23.8 days in group B, p=0.97). A similar result was obtained in the ITT analysis.

**Table 2.** Primary and secondary endpoints analyses comparing secukinumab + SoC (group A) to SoC alone (group B).

| Group | A (SEC + SoC)<br>n=25 | B (SoC alone)<br>n=23 | p value |
| --- | --- | --- | --- |
| **Primary endpoint** |  |  |  |
| VFD-28 (days), mean (SD) | 23.7 (9.9) | 23.8 (9.6) | 0.62 |
| **Secondary endpoints** |  |  |  |
| Mechanical Ventilation, n (%) | 5.0 (20) | 5.0 (22) | 1.00 |
| Hospital days, mean (SD) | 9.3 (8.1) | 11.0 (7.4) | 0.34 |
| ICU, n (%) | 7 (28) | 9 (39) | 0.54 |
| ICU days, mean (SD) | 4.0 (9.0) | 3.2 (6.3) | 0.55 |
| WHO scale $\leq 1$, n (%) | 12 (48) | 6 (26) | 0.14 |
| WHO scale, mean (SD) | 2.3 (2.1) | 2.1 (1.5) | 0.31 |

VFD-18, number of ventilator-free days after 28 days of follow-up; ICU, intensive care unit; WHO scale, World Health Organization Ordinal Scale for Clinical Improvement.

The analysis of secondary endpoints showed no significant differences in all evaluated parameters. Although there was an appearant decrease in ICU admissions (28% *vs*. 39%) and hospitalization length (9.3 *vs*. 11.0 days) in secukinumab-treated patients than in the control group, these differences were not statistically different (Table 2). Similarly, there was an nominally longer duration of mechanical ventilation (3.8 *vs*. 2.0 days) and longer permanence in the ICU (4.0 *vs*. 3.2 days) in secukinumab-treated than control patients, but this was also without statistical significance.

The frequency of serious adverse events (SAE) was generally comparable in both groups (20% *vs*. 26%; p=0.73) (Table 3). Secukinumab-treated individuals had an appearant increase in the rate of secondary bacterial infection (16% *vs*. 9%; p=0.7) and circulatory shock (16% *vs*. 4%; p=0.3), albeit these were not significantly different between groups. The incidence of pulmonary thromboembolism (PTE) was lower in the secukinumab-treated than in the control group (4% *vs*. 26%; p=0.04). The incidence of PTE (per 100 patient-days) was 0.14 in group A and 0.93 in group B, with a relative risk reduction of 85% (Table 3). There was one death and one patient who developed acute kidney injury (KDIGO III) requiring hemodialysis in each group. There was only one fungal infection (vaginal candidiasis, treated with oral antifungal, without study interruption) in secukinumab-treated patients and none in the control group. No allergic reactions or reactions at the injection site occurred in either study arm.

**Table 3.** Incidence of adverse events of interest in secukinumab + standard of care (group A) compared to standard of care alone (group B).

| Group | A (SEC + SoC)<br>n=25 | B (SoC alone)<br>n=23 | p value |
| --- | --- | --- | --- |
| Patients with SAE, n (%) | 5 (20) | 6 (26) | 0.73 |
| Any SAE, n (Incidence/100 patient-days) | 16 (2.3) | 11 (1.7) | 0.56 |
| Pulmonary thromboembolism, n (%) | 1 (4) | 6 (26) | 0.04* |
| Secondary bacterial infection, n (%) | 4 (16) | 2 (8.7) | 0.67 |
| Secondary fungal infection, n (%) | 1 (4) | 0 | 1.00 |
| Circulatory shock, n (%) | 4 (16) | 1 (4) | 0.35 |
| Acute kidney injury KDIGO III, n (%) | 1 (4) | 1 (4) | 1.00 |
| Death, n (%) | 1 (4) | 1 (4) | 1.00 |

SAE, serious adverse events; \* $p < 0.05$ = statistically significant.

Chronological analysis (from day-0 to day-14) of the neutrophil/lymphocyte ratio (NLR), ferritin, and CRP levels during hospitalization revealed a very similar evolution of the inflammatory markers. NLR tended to increase from days 10 to 14 in group A and decrease in group B. Still, there were few hospitalized patients during this period, and the differences were not statistically significant (Figure 2).

**Figure 1.**
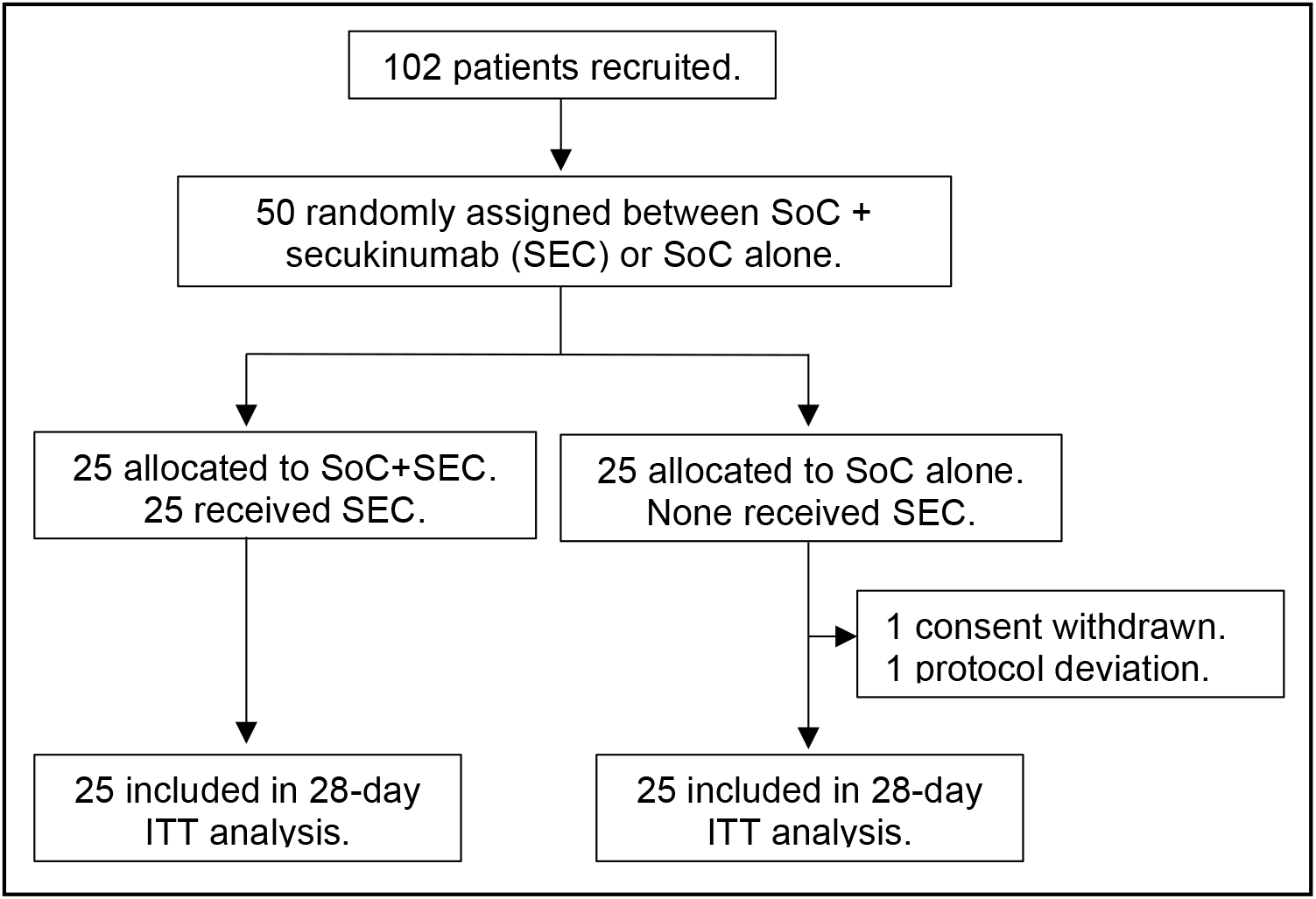
Trial profile. SoC, standard of care.

**Figure 2.**
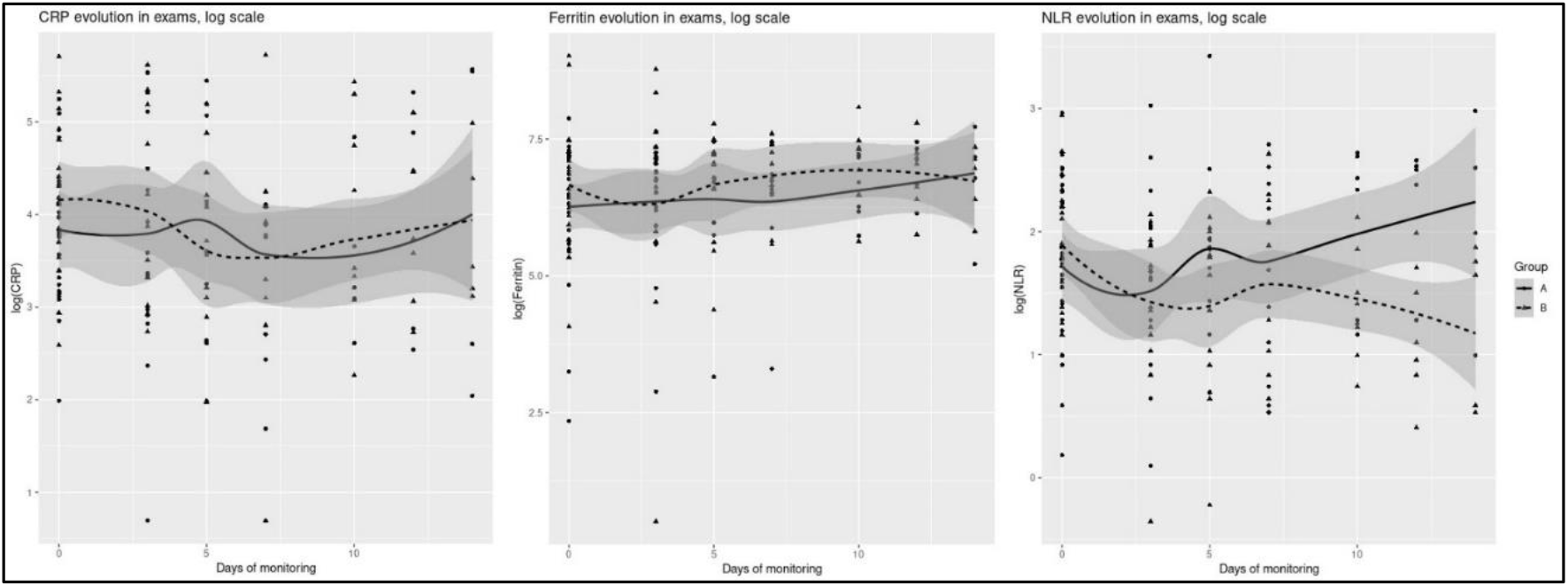
Chronological analysis of blood tests. CRP, C-reactive protein; NLR, neutrophil-lymphocyte ratio. Shadow areas represent the 95% confidence interval.

The WHO Ordinal Scale for Clinical Improvement had comparable evolution, from randomization to day 28 in both groups, as seen in Figure 3. The score in the WHO Ordinal Scale for Clinical Improvement at day 28 was not different between groups (2.3 points in group A *vs*. 2.1 in group B). There was a higher proportion of patients in group A (48%) than group B (26%), achieving a score of one on the scale throughout the 28 days of follow-up, although without significance (p=0.14), as shown in Figure 3

**Figure 3.**
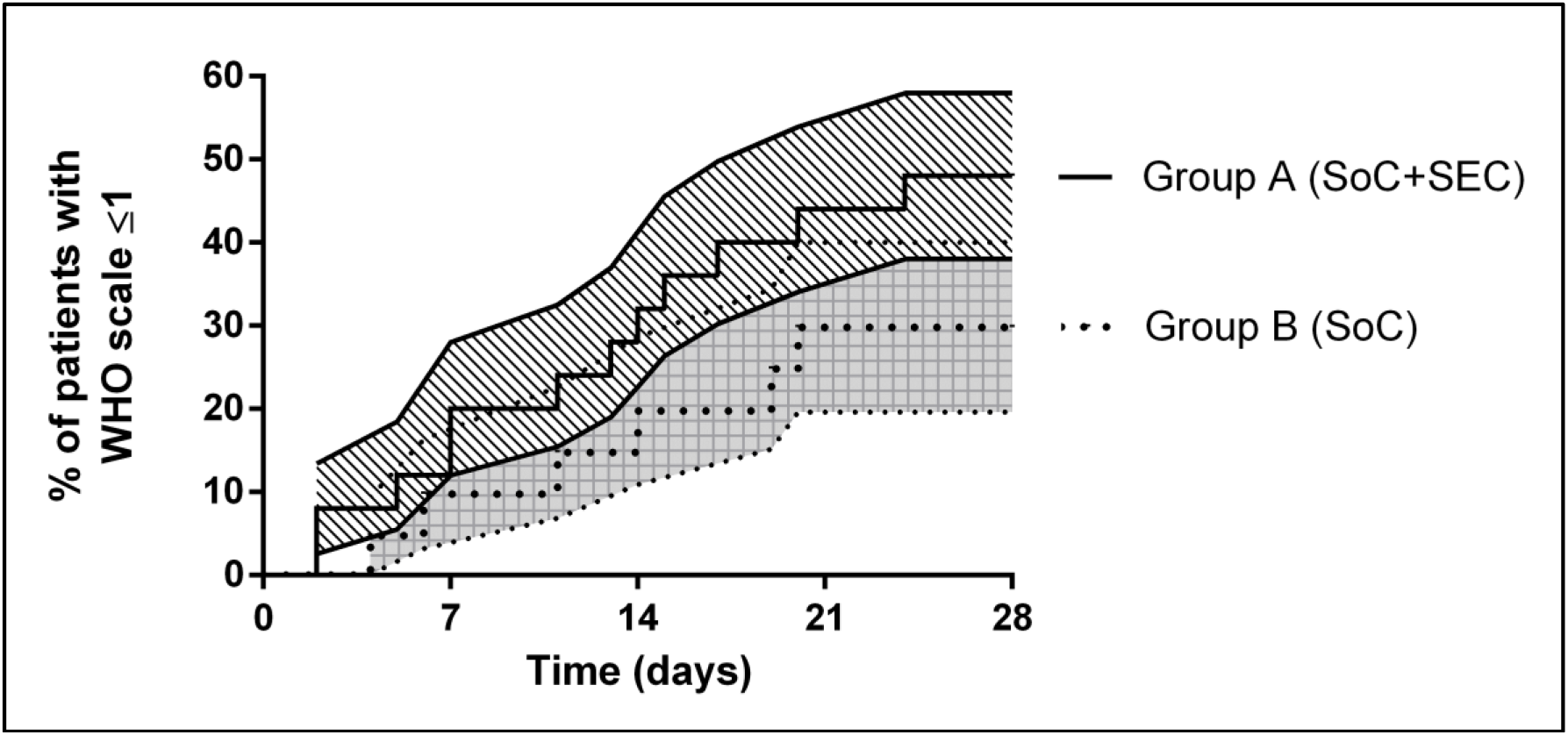
Cumulative proportion of patients who reached World Health Organization (WHO) Ordinal Scale for Clinical Improvement evolution ≤ 1 from day 0 to day 28. Error lines (shadow areas) represent the standard error (SE).

Viral clearance, as defined by the fold change (2-ΔΔCT) in RNA viral load in the upper airway between day-0 and day-7, was similar in both groups. After seven days, there was a decrease of 83% in group A and 77% in group B, as shown in Figure 4.

**Figure 4.**
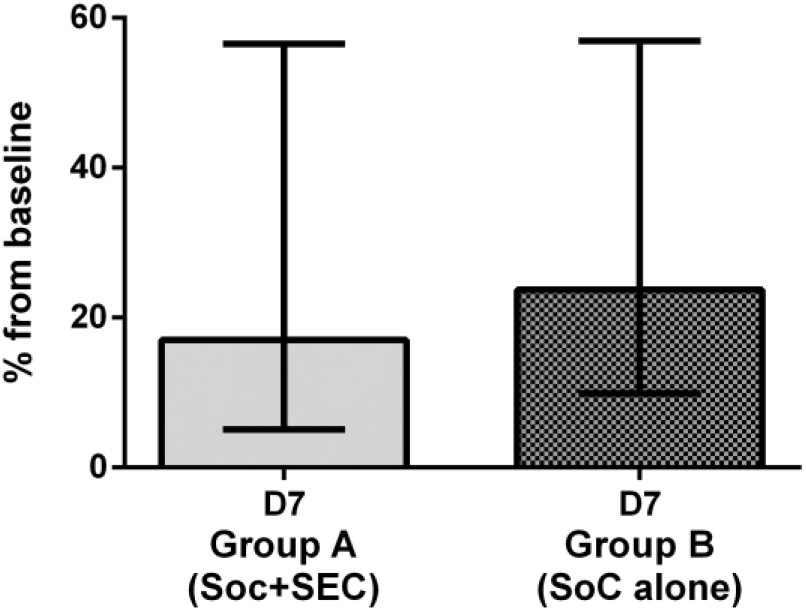
Viral clearance in upper airways from randomization to day 7. Error bars represent the 95% confidence interval of the mean fold change.

## 4. DISCUSSION

To the best of our knowledge, this is the first randomized clinical trial exploring the effects of secukinumab in COVID-19 patients. Our results show that secukinumab was well tolerated and not associated with a significant increase of adverse events or secondary infections. However, the anti-IL-17 antibody was not effective in ameliorating the pulmonary failure observed in severe acute respiratory syndrome COVID-19 patients. Indeed, there was no decrease in the number of ventilator-free days at day 28 with secukinumab in addition to the standard therapy used. Moreover, days in the hospital, admission to ICU, and time to recovery (as assessed by the WHO score) were similar in both groups. Of special interest, our study showed that the number of clinically relevant thromboembolic events was significantly lower in secukinumab-treated patients.

As seen in Table 2, there was no difference in the percentage of individuals intubated and the duration of intubation. Similarly, ICU admission and ICU stay were similar between both groups. These results suggest that the most severe forms of the disease are not altered in any significant way by anti-IL-17 treatment. On the other hand, there were parameters that, albeit not significant, showed a potential for secukinumab in ameliorating the less severe forms of the disease. Indeed, there was a two-day difference in hospitalization, mainly because there was an early amelioration of WHO scores in secukinumab-treated patients.

Our study showed no difference between groups in the incidence of adverse events, and no unexpected events were observed in this small population (n= 25 of patients treated with secukinumab). These findings are consistent with published data showing no increased risk of COVID-19 infection or its complications in patients taking IL-17 inhibitors for immune-mediated diseases ^(23-25)^. There was a trend for more infections in secukinumab-treated individuals, but there was no statistical difference between the two groups. The tendency to increase NLR as the disease progressed seen in group A (Figure 2) could reflect that difference since neutrophil blood count started to rise on day-7 (data not shown).

It is estimated that 1% of patients tested for SARS-CoV-2 develop pulmonary thromboembolism (PTE) ^(26)^. Clinically relevant PTE was estimated at 8% (95%CI, 4–11%) ^(27)^. Notably, in critically ill patients, this prevalence is remarkably higher, with prevalences reported of up to 76% in this population ^(28)^. In our cohort, the prevalence of PTE in the control group (26%) was close to that reported in the literature. Therefore, the substantial reduction of pulmonary thromboembolism (PTE) seen in secukinumab-treated patients was remarkable. There is evidence that IL-17 may contribute to thromboinflammation under various circumstances. Upon stimulation, COVID-19 platelets released significantly larger amounts of IL-17, among other cytokines, compared with control platelets ^(29)^. IL-17A promotes in vitro platelet aggregation and activation, neutrophil extracellular trap (NET) release, and vein endothelial cell activation. These effects were lost in platelets deficient for the IL-17 receptor (IL-17R) ^(30-32)^, resulting in less deep venous thrombosis (DVT) formation in two animal models ^(33, 34)^. Besides, a series of thrombectomies observed that the coronary thrombus, superimposed on ruptured atherosclerotic plaques, contains NETs coated with IL-17 ^(35)^. While this six-fold reduction of clinically significant PTE in COVID-19 patients treated with secukinumab is of interest, larger studies are needed to confirm this protective effect of blocking IL-17.

This study had some limitations. It was open-label with a small sample size. The sample size was calculated based on the risk of intubation and death observed at the beginning of the epidemic in Brazil. When inclusions started in September 2020, all patients were already taking corticosteroids because of its benefits in oxygen-dependent COVID-19 patients ^(36)^. Possibly, this issue improved the challenge to show the significant effects of an anti-inflammatory intervention on top of steroid use. In fact, smaller studies with another anti-cytokine therapy (anti-IL-6R) have also reported no improvement in COVID-19 patients, while two larger trials clearly showed a benefit ^(37)^. Furthermore, the optimal dose, route of administration, and timing of secukinumab to ensure therapeutic tissue concentrations in COVID-19 are unknown. The dose regimen chosen in this study was the maximum weekly dose, currently approved for other indications.

In conclusion, secukinumab exhibited a satisfactory safety profile in severe COVID-19 patients. Although the blockade of anti-IL-17A with secukinumab appears not to show a clear benefit, compared to the standard of care, in the context of COVID-19 pulmonary failure, there was a significant reduction of pulmonary thromboembolism. Larger clinical trials of secukinumab in COVID-19 may be warranted.

## Data Availability

De-identified data can be made available to others on approval of a written reasonable request to the corresponding author.

## Contributors

GGR, MMT and SQL accessed and verified the data. GGR and MMT contributed to the literature search, study design, and data interpretation. AJAO, ADCS, ATNS, HCG, ICG, SQL did all study assessments, study visits, and completed data entry. DSG supported the local laboratory assessments and AFM provided the local pharmaceutical assistance. RSA and RPS were responsible for all the central RT-PCR tests. JSSBF was the independent statistician and analyzed all data. FF designed the electronic case report form (CRF) and did the block randomization. GGR, HCG, MMT, FF, RT, RCL, RSA, RPS, SQL cooperate with critical revision of the manuscript. All authors contributed to the writing of the article and approved its submission. GGR and MMT were responsible for the decision to submit the article.

